# Utility of Lung Ultrasound in COVID-19: A Systematic Scoping Review

**DOI:** 10.1101/2020.06.15.20130344

**Authors:** Michael M Trauer, Ashley Matthies, Nick Mani, Cian Mcdermott, Robert Jarman

## Abstract

Lung ultrasound (LUS) has an established evidence base and has proven useful in previous viral epidemics. An understanding of the utility of LUS in COVID-19 is crucial to determine its most suitable role based on local circumstances. A scoping review was thus undertaken to explore the utility of LUS in COVID-19 and guide future research.

33 studies were identified which represent a rapidly expanding evidence base for LUS in COVID-19. The quality of the included studies was relatively low. However LUS appears to be a highly sensitive and fairly specific test for COVID-19 in all ages and in pregnancy and is almost certainly more sensitive than CXR. The precise diagnostic accuracy of LUS may be influenced by various factors including disease severity, pre-existing lung disease, scanning protocol, operator experience, disease prevalence and the reference standard.

High quality research is needed in various fields including: diagnostic accuracy in undifferentiated patients; triage and prognostication; monitoring progression and guiding interventions; persistence of residual LUS findings; inter-observer agreement; and the role of contrast-enhanced LUS.

## INTRODUCTION

### Rationale

Coronavirus Disease 2019 (COVID-19) is caused by Severe Acute Respiratory Syndrome Coronavirus 2 (SARS-CoV-2) and was declared a global pandemic on the 11^th^ March 2020 by the World Health Organisation. As of the 10^th^ of June, there have been over seven million confirmed cases and over 400,000 deaths [^1^].

The evidence base for lung ultrasound (LUS) is well established. In 2008, LUS was found to have an accuracy of greater than 90% for some of the most common causes of dyspnoea [^2^]. In 2011, an international, evidence-based, consensus statement recommended its use in pneumothorax, interstitial syndrome, consolidation and effusion [^3^]. In 2015, a prospective study of over a thousand patients found incorporation of LUS into clinical assessment significantly improved sensitivity (97%) and specificity (97.4%) for acute heart failure [^4^]. And in 2018, a meta-analysis of over five thousand patients found LUS to be 92% sensitive and 93% specific for community-acquired pneumonia [^5^].

LUS has also proven useful during recent viral epidemics. In the 2009 influenza (H1N1) epidemic, LUS was found to be accurate in differentiating viral and bacterial pneumonia [^6^], and during the avian influenza (H7N9) epidemics LUS was found to be superior to CXR (sensitivity 94%, specificity of 89%) [^7,8^].

In admitted patients with COVID-19, CXR has a reported sensitivity of between 59% and 69% [^9,10^]. In ambulatory patients with symptomatic COVID-19, CXR sensitivity has been reported at 42% [^11^].

The sensitivity of PCR for COVID-19 has been estimated at 70% [^12^] and depends upon factors including the quality of sampling and stage of illness.

CT of the thorax is highly sensitive for COVID-19 [^13^] however LUS has several logistical advantages. The capacity to perform routine CT in suspected COVID-19 may become overwhelmed if large numbers of patients attend hospital. LUS has also been shown to reduce healthcare worker exposure to COVID-19 by reducing the intra-hospital transfers associated with conventional imaging [^14^]. Other advantages of LUS over CT include reduced cost, repeatability, lack of radiation exposure and rapid image acquisition time [^15^].

LUS has been shown to improve diagnostic accuracy in patients who present with acute respiratory symptoms [^16^] and is increasingly used by frontline clinicians who assess these patients. Ultrasound machines continue to improve in quality, affordability and portability [^17^] and new technologies such remote teleguidance have the potential to further extend the accessibility of point-of-care ultrasound.

The LUS findings in COVID-19 are well described and include B lines, pleural line abnormalities and consolidation [^18^]. However the most suitable role for LUS in COVID-19 is still unclear. Various roles have been proposed including triage, diagnosis, prognostication, severity scoring, monitoring progression, and guiding interventions [^19^]. An understanding of the utility of LUS is crucial to determine its most suitable role in COVID-19 based on local circumstances.

### Objectives

To review the evidence of the utility of LUS in COVID-19 and guide future research

- Population: Patients with suspected or confirmed COVID-19
- Concept: The utility of LUS
- Context: Clinical management

## METHODS

### Protocol and registration

The protocol was drafted in line with PRISMA [^20^] and registered on https://figshare.com/ on 13/6/2020 (10.6084/m9.figshare.12478820)

### Inclusion criteria

- Patients of any age with suspected or confirmed COVID-19
- Explores the utility of LUS in COVID-19
- Trials or case series (prospective or retrospective)

### Exclusion criteria

- Case reports and recommendations
- Non-English language

### Information sources

Traditional online databases were searched including: Medline, Embase, SCOPUS, The Cochrane Library, The TRIP database, Google Scholar and www.clinicaltrials.gov.

Given the dynamic nature of the pandemic, other less traditional sources were also searched including point-of-care ultrasound (PoCUS) websites, specialty college websites, pre-publication websites and social media platforms (see Appendix I).

### Search strategy

An initial search strategy was formulated by MT and reviewed by AM using the PRESS checklist [^21^]. This initial search was performed on two databases (Medline and Embase) (See Appendix II).

Keywords were identified from the above abstracts and another search was performed of all relevant databases (See Appendix III).

### Selection of sources

A screening and selection tool was applied to the identified studies by two independent reviewers (MT & AM) with a third reviewer (NM) available to resolve disagreements (See Appendix IV).

The reference lists from these included studies were then reviewed for further relevant studies. The authors of the included studies were contacted regarding relevant unpublished or recently published evidence.

### Data items

Data was extracted on study design, numbers of participants, population, and data relating to the utility of LUS in COVID-19.

### Synthesis of results

Given the heterogeneity of the data, findings are described in a narrative style.

## RESULTS

### Selection of sources of evidence

A flow diagram in line with PRISMA [^20^] is presented in Figure 1 and displays the number of studies screened, excluded and assessed.

**Figure 1.**
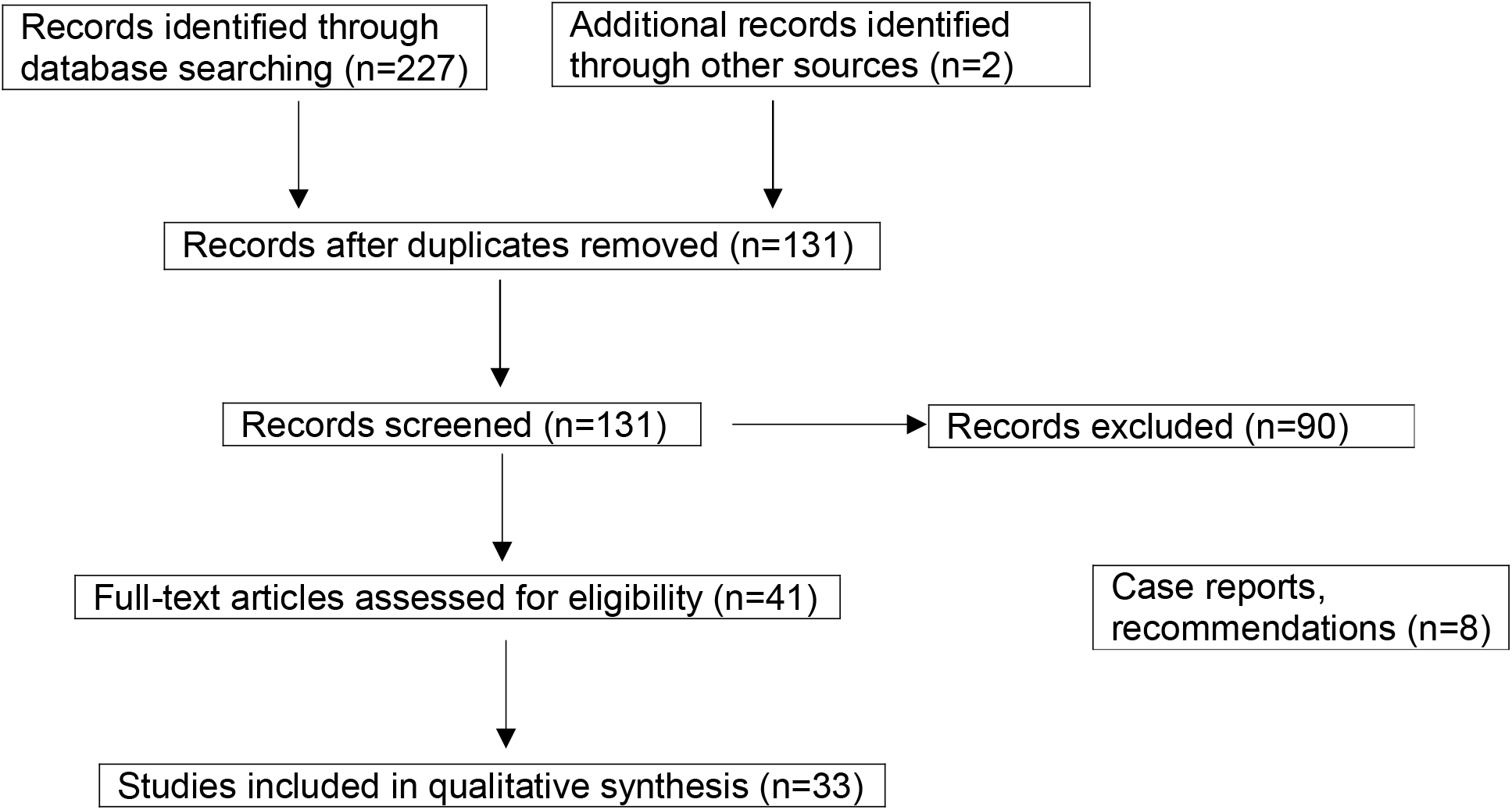
PRISMA Flow diagram.

### Characteristics of sources of evidence

A total of 33 studies were identified from countries including China, Italy, Spain, France and the USA. 17 were single-centre, three were multi-centre and the numbers of participants in each study ranged from three to 107.

The topics explored in each study are summarised in Table 1. The characteristics of each included study are summarised in Appendix V.

**Table 1.**

| Topic explored | Studies identified |  |
| --- | --- | --- |
| Description of LUS findings | 5 |  |
| Comparison of LUS to a reference standard | 9 | 7 vs CT and 2 vs PCR |
| Serial LUS imaging | 2 |  |
| Technological innovation | 4 |  |
| Special groups | 7 | 3 in pregnancy and 4 in children |
| Reviews | 6 | 1 meta-analysis and 5 narrative |
| Total | 33 |  |

### Studies that describe LUS findings in COVID-19

The LUS findings in COVID-19 have already been well described and consist of B lines, pleural line abnormalities and consolidations usually without pleural effusion [^18, 22, 23, 24, 25^]. Mohamed *et al*. [^26^] performed a meta-analysis of such studies (seven studies, 122 patients) and found B lines were the most common and consistent finding but other LUS findings had a high degree of heterogeneity.

### Comparison of LUS to a reference standard (CT)

In non-peer reviewed data from Tung-Chen *et al*. [^27^], 51 adults presented to ED with confirmed or suspected COVID-19, both received CT and LUS and 67% were admitted. LUS was performed by a single, experienced operator blinded to CT and clinical findings. CT was suggestive of COVID-19 in 37 patients and all 37 were identified by LUS (sensitivity 100%, specificity 79%). The area under the ROC curve was greater for LUS (86%) than for PCR (63%) for detecting CT abnormalities.

In non-peer reviewed data from Hankins *et al*. [^28^], 49 patients over the age of 14 and without underlying lung disease attended ED with symptoms of COVID-19. Compared to CT, LUS had a sensitivity and specificity of 100% and 80% respectively when performed by the treating clinician however this fell to 92% and 37% when the images were reviewed in isolation by an independent clinician. Compared to CT, the sensitivity of CXR and crackles on auscultation was 25% and 8% respectively.

In Yang *et al*. [^29^], 29 adult patients with confirmed COVID-19 received simultaneous LUS and CT. The lung fields were divided into 12 regions: 63% of regions displayed abnormal findings on LUS (3 or more B lines, consolidation or pleural effusion) compared to 39% on CT (ground-glass opacity, consolidation or pleural effusion). The authors concluded that LUS was more sensitive than CT at identifying the above COVID-19 findings.

In non-peer reviewed data from Benchoufi *et al*. [^30^], 107 adult patients attended ED with confirmed or suspected COVID-19, all received both CT and LUS and CT was typical for COVID-19 in 80%. When LUS was considered as a four-category ordinal scale of severity, there was moderate agreement with CT, kappa 0.52 (0.38-0.66), however when this was reduced to a binary outcome (normal vs pathologic) there was strong correlation (sensitivity 95%, specificity 83%).

In Lu *et al*. [^31^], 30 adult patients admitted with confirmed COVID-19 received both LUS and CT. The ability of LUS to predict the severity of COVID-19 using CT as the reference standard was assessed. The diagnostic accuracy of LUS for no, mild, moderate and severe disease was 93%, 77%, 77% and 93% respectively.

In Poggliali *et al*. [^32^] and Lyu *et al*. [^33^], a total of 20 adult patients received both LUS and CT and the authors noted a strong correlation between LUS and CT.

### Comparison of LUS to a reference standard (PCR)

In Peyrony *et al*. [^34^], 47 patients presented to an Emergency Department with suspected COVID-19 and received LUS. The presence of bilateral B lines had a sensitivity and specificity of 77% and 89% respectively.

In Bar *et al*. [^35^], 100 adults presented to an Emergency Department with suspected COVID-19. 31% were PCR positive but CT results were not recorded. The combination of qSOFA (quick sequential organ failure assessment) score and LUS gave an area under the ROC curve of 0.82 with sensitivity and specificity of 97% and 62% respectively.

### Special groups

Four paediatric studies were identified comprising 26 patients, all admitted with confirmed COVID-19 and ranging from neonatal to 15 years of age. In Musolino *et al*. [^36^], Feng *et al*. [37] and Gregorio-Hernandez *et al*. [^38^], all 18 patients (ten children and eight neonates) demonstrated LUS findings. In Denina *et al*. [^39^], five of the eight children demonstrated LUS findings (one of four children with mild disease but all four cases of moderate to severe disease).

Three obstetric studies were identified comprising 16 patients, all admitted with confirmed COVID-19. In Buonsenso *et al*. [^40^] and Giannini *et al*. [^41^], all eight patients demonstrated typical LUS findings. In Yassa *et al*. [^42^], seven of the eight women demonstrated LUS findings and LUS changed clinical management in 87.5% of cases.

## DISCUSSION

### Methodology

There were various issues regarding the methodology of the included studies including convenience sampling, unrepresentative populations (often only admitted patients), lack of power calculations, variability of index test (operator experience, scanning protocol), variability of reference standard (CT, single PCR test, multiple PCR tests) and reproducibility. A summary of the levels of evidence of the included studies according to the Oxford centre for evidence-based medicine [^43^] is displayed in table 2.

**Table 2.**

| Level of Evidence | Number of studies |
| --- | --- |
| 1a | 0 |
| 1b | 0 |
| 1c | 0 |
| 2a | 0 |
| 2b | 3 |
| 2c | 0 |
| 3a | 6 |
| 3b | 20 |
| 4 | 3 |
| 5 | 1 |

### Comparison of LUS to a reference standard

CT is highly sensitive for COVID-19 [^13^] and therefore is generally assumed to be the reference standard for LUS. However this assumption was challenged by Yang *et al*. [^29^] and Feng *et al*. [^37^] who concluded that LUS may in fact be more sensitive than CT.

The included studies are suggestive that LUS is highly sensitive for COVID-19. However sensitivity may be affected by factors including disease severity and scanning technique. Lu *et al*. [^15^] and Denina *et al*. [^39^] both found LUS to have greater sensitivity in more severe disease. Regarding LUS technique, various protocols have been suggested ranging from a limited scan of just the anterolateral zones to a comprehensive ‘lawn-mower’ technique where the probe is slid along each intercostal space [^44^]. Given the disease is known to have a patchy distribution [^18^] it would be plausible that a more comprehensive protocol would be more sensitive however there is also evidence that LUS findings do not depend on the number of zones assessed [^45^].

PCR testing (in its current form) is highly specific but relatively insensitive [^12^]. LUS specificity may therefore be under-estimated in studies where LUS is compared only to PCR. For example in Bar *et al*. [^35^] specificity was 62% (vs PCR) but in Tung-Chen *et al*. [^27^] specificity was 79% (vs CT).

Operator experience may also affect specificity as an expert will be able to better correlate different LUS patterns with different disease processes. In Benchoufi *et al*. [^30^], specificity was 83% however this was based on the LUS being simply normal or pathologic. In Peyrony *et al*. [^34^] specificity was 89% however this was based on only the presence of bilateral B lines. A more nuanced LUS assessment may lead to greater specificity.

There may be particular LUS findings and patterns that are more specific for COVID-19. In Volpicelli *et al*. [^46^] the authors described a LUS artefact called ‘light beam’, defined as a broad, lucent, band-shaped, vertical artefact moving rapidly with sliding and arising from a regular pleural line. The authors stated that in a series of 100 patients (unpublished data) this finding was present in 48 of 49 patients with confirmed COVID-19 but in none of 12 patients with negative swabs and alternative diagnoses. Futhermore, Soldati *et al*. [^47^] argued that relative specificity can be attributed to a bilateral, patchy distribution with spared areas and multifocal confluent B lines (‘white lung’), especially in relatively young patients without a history of lung disease.

There is a spectrum of LUS findings in COVID-19 ranging from subtle to highly suggestive. Many studies have focused on LUS severity scores however in terms of diagnostic utility an assessment of likelihood rather than severity may be more useful. The differential diagnosis of each specific patient will also influence which LUS findings are most specific to COVID-19. If the alternative is pulmonary oedema, the presence of pleural thickening and irregularity is relatively specific for COVID-19. However if the alternative is pulmonary fibrosis this finding would not help to discriminate between these processes. In Hankins *et al*. [^28^], diagnostic accuracy was higher when LUS was interpreted by the treating clinician as opposed to being reviewed in isolation. This highlights the importance of integrating LUS findings with clinical findings. This Bayesian approach of combining a pre-test probability with point-of-care ultrasound findings is well described [^48^].

It should be noted that all of the included studies were conducted during a period of high disease prevalence and it is likely that measures of diagnostic accuracy will be affected by fluctuations in disease prevalence over time [^49^].

### Serial LUS imaging

In Xing *et al*. [^50^], 20 adult patients with confirmed COVID-19 underwent 36 scans at various time intervals after onset of symptoms. The authors found that the extent of LUS findings reached a peak at the second week and then there was gradual improvement (but not complete resolution) until the fourth week.

In Shkoohi *et al*. [^51^], three physicians with confirmed COVID-19 monitored themselves at home and in all cases the LUS findings had resolved by day 14.

More information is urgently needed regarding the persistence of LUS findings as clinicians will be increasingly encountering patients who may have recently recovered from COVID-19.

### Inter-observer agreement

Good inter-observer agreement of LUS findings was found between experts [^35^] and between experts and novices [^30^]. However it was noted that removal of the practical element of novice training significantly reduced inter-observer agreement [^28^]. It was also noted that inter-observer agreement varied between different LUS findings, being highest for consolidation and lowest for pleural thickening [^28^].

Inter-observer agreement will depend on the extent of training the novice has received and a wide array of training protocols have been described. In Benchoufi *et al*. [^30^], only 30 minutes of theoretical and 30 minutes of practical training was required. However it has previously been suggested that 25 scans are necessary to achieve competency in LUS [^52^].

Further studies relating to inter-observer agreement are warranted however it appears the element of practical training is important. Novel technologies such as remote teleguidance could help to achieve this.

### Technological innovation in LUS

New technologies may play an important role in augmenting the potential utility of LUS in COVID-19. Several avenues are currently being explored including artificial intelligence, deep learning, robotic LUS and contrast-enhanced LUS.

Dong *et al*. [^53^] stated that artificial intelligence or other quantitative image analysis methods were urgently needed to maximise the value of imaging modalities including LUS.

Roy *et al*. [^54^] created a deep model of automatic analysis from an annotated LUS data set and noted that this achieved ‘satisfactory results’ on all tasks including predicting disease severity.

Evans *et al*. [^55^] noted that robotic ultrasound equipment is being used at Zhejiang Provincal People’s Hospital in Hangzhou, China.

In Soldati *et al*. [^56^], three patients with confirmed COVID-19 received contrast-enhanced ultrasound (CEUS). Perfusion defects were noted within these lesions and the authors concluded that this was at least in part caused by ischaemic or necrotic changes rather than inflammation or atelectasis. This is consistent with the findings of Huang *et al*. [^18^] who noted the lack of colour Doppler signal within subpleural consolidations in COVID-19. If these peripheral lung lesions are in fact infarcts this may have major implications for clinical management and therefore this question deserves further attention.

### Special groups

The issue of ionizing radiation is of great concern in children and pregnant women. Several small studies were identified that examined the utility of LUS in COVID-19 in these patient groups and were suggestive that LUS is as useful as it is in non-pregnant adults.

### Limitations

The recent emergence and dynamic nature of the COVID-19 pandemic has led to the rapid publication of research and it is inevitable that new studies will continue to be released before this review is published.

A thorough and systematic literature search was performed including non-traditional sources (see Appendix I) however all relevant evidence may not have been identified due to publication bias and non-English language publications being excluded.

## Conclusion

The evidence base for LUS in COVID-19 is rapidly expanding but the methodological quality of the identified studies was relatively low.

It is difficult to make a precise estimate of diagnostic accuracy of LUS in COVID-19 as both sensitivity and specificity may be influenced by various factors including disease severity, pre-existing lung disease, scanning protocol, operator experience, disease prevalence and the reference standard. However, LUS appears to be a highly sensitive and fairly specific test for COVID-19 in all ages and in pregnancy. LUS is almost certainly more sensitive than CXR for COVID-19 and possibly more sensitive than CT.

High quality research is needed to better define the utility of LUS in COVID-19 and thus inform clinicians of its most suitable role in a local context. Although the LUS findings in COVID-19 are now well described, further research is needed regarding the relative specificity of the various LUS findings and patterns. High quality, prospective studies assessing diagnostic accuracy in undifferentiated patients in an era of lower prevalence would also be of great value. The role of LUS in triage, prognostication, severity scoring, monitoring progression and guiding interventions has not yet been adequately explored. An understanding of the persistence of residual LUS findings post infection will be increasingly important going forwards. Larger studies assessing inter-observer agreement would both estimate reproducibility but may also help inform necessary training standards for novices. Further research into contrast-enhanced LUS and colour Doppler is warranted as this may significantly augment traditional LUS and contribute to a broader understanding of the disease process. International consensus is required regarding training standards, scanning protocols and an appropriate reference standard.

## Data Availability

REVIEW ARTICLE SO NOT APPLICABLE

## Appendix I. Extended literature search

Specialised point-of-care ultrasound websites

- Zedu Ultrasound Training Solutions: https://www.ultrasoundtraining.com.au/news/covid-19-pocus-resources
- Ultrasound G.E.L. Podcast: Gathering Evidence from the Literature: https://www.ultrasoundgel.org/articles

Specialty college websites

- Royal College of Emergency Medicine: https://www.rcemlearning.co.uk/research/
- Intensive Care Society: https://ics.ac.uk/ICS/ICS/FUSIC/FUSIC_COVID-19.aspx

Pre-publication websites

- MedRxiv, The preprint server for health sciences: https://www.medrxiv.org
- Figshare: https://figshare.com/browse

Social media

- Twitter hashtag: #pocusforcovid

## Appendix II. Initial search

Databases to be searched

1. Ovid MEDLINE ^®^ to June 13^th^ 2020
2. Embase 1974 to 2020 June 13^th^ 2020

Search strategy:

1. Lung OR chest OR thorax OR thoracic
2. Ultrasound OR ultrasonography OR sonography
3. COVID OR COVID-19 OR coronavirus OR SARS-CoV 2
4. 1 AND 2 AND 3
5. 4 AND remove duplicates

## Appendix III. Second search

Databases to be searched

1. Ovid MEDLINE ^®^ and Epub Ahead of Print, In-Process and Other Non-Indexed Citations, Daily and Versions 1946 to June 13^th^ 2020
2. Embase 1974 to 2020 June 13^th^
3. Scopus
4. The Cochrane Library
5. The TRIP database
6. Google Scholar
7. www.clinicaltrials.gov
8. JBI Database of Systematic Reviews and Implementation Reports
9. Cochrane Database of Systematic Reviews
10. Cumulative Index to Nursing and Allied Health Literature (CINAHL)
11. Evidence for Policy and Practice Information (EPPI)
12. Epistemonikos

Second search strategy

1. Lung OR chest OR thorax OR thoracic OR pulmonary

1. Ultrasound OR ultrasonography OR sonography OR ultrasonic
2. COVID OR COVID-19 OR coronavirus OR SARS
3. 1 AND 2 AND 3
4. 5 AND remove duplicates

## Appendix IV. Screening and selection tool

| LUS in COVID-19: Screening and Selection Tool |  |  |
| --- | --- | --- |
| Reviewer name: |  | Date: |
| Title: |  |  |
| Author name: | Year: | Journal: |
|  | Inclusion Criteria | Exclusion Criteria |
| Diagnostic test: | ☐ LUS |  |
| Population: | ☐ Patients of any age with suspected or confirmed COVID-19 |  |
| Concept: | ☐ Utility of LUS |  |
| Context: | ☐ Clinical management of COVID-19 |  |
| Study design: | ☐ Case series<br>☐ Experimental or observational studies<br>☐ Reviews | ☐ Case reports<br>☐ Recommendations |
| Overall decision: | ☐ Included | ☐ Excluded |

## Appendix V. Study Characteristics and Results

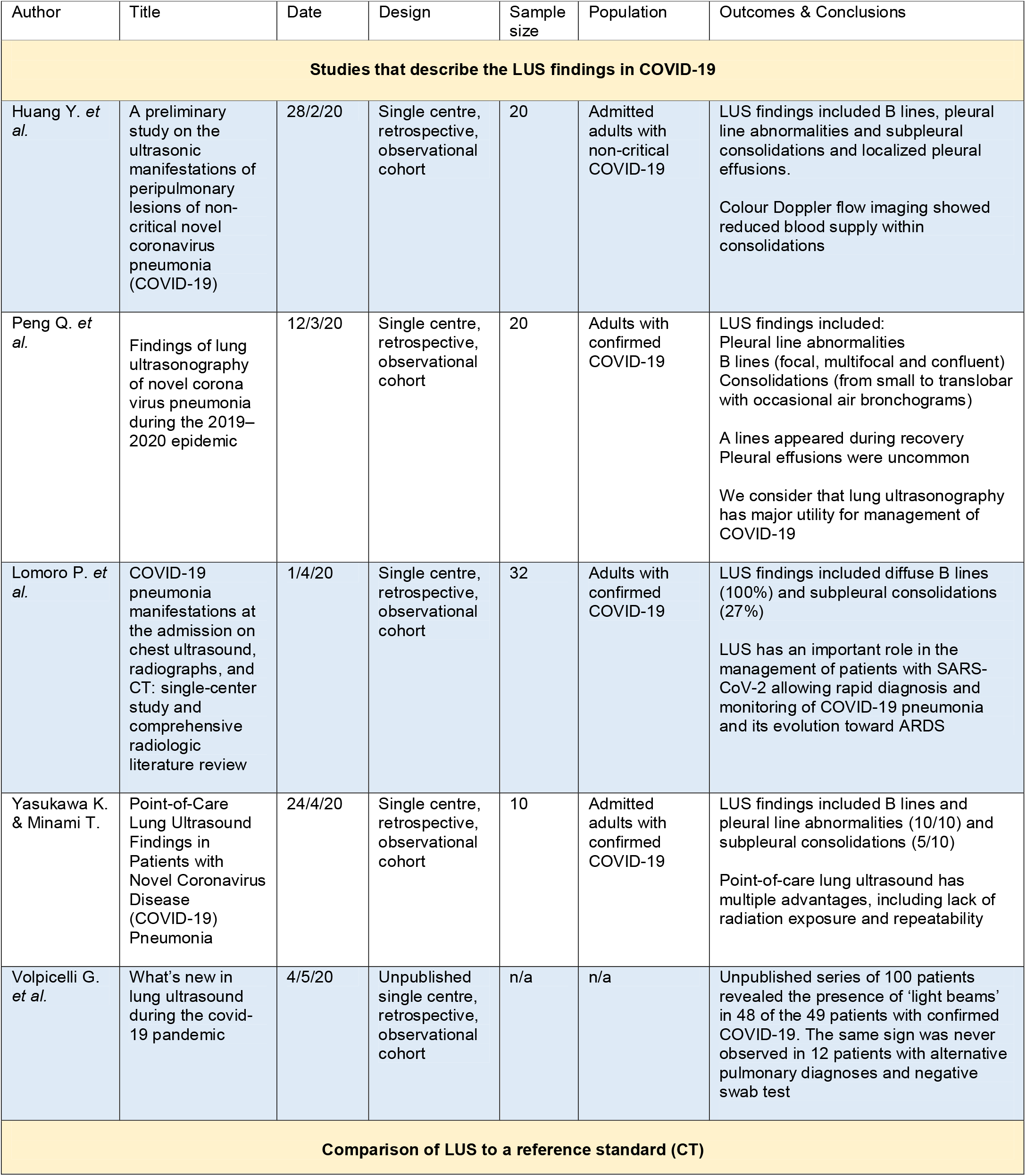
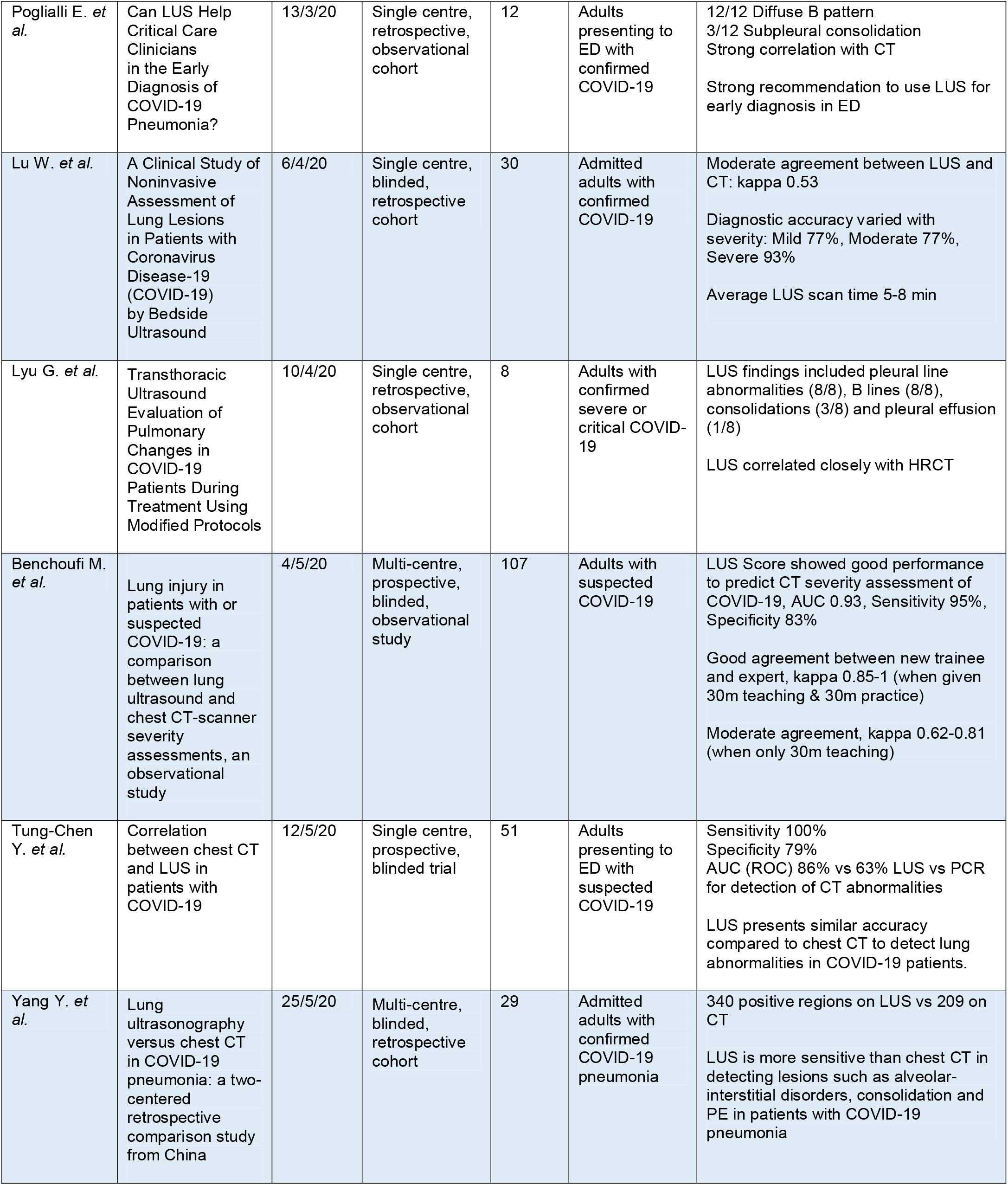
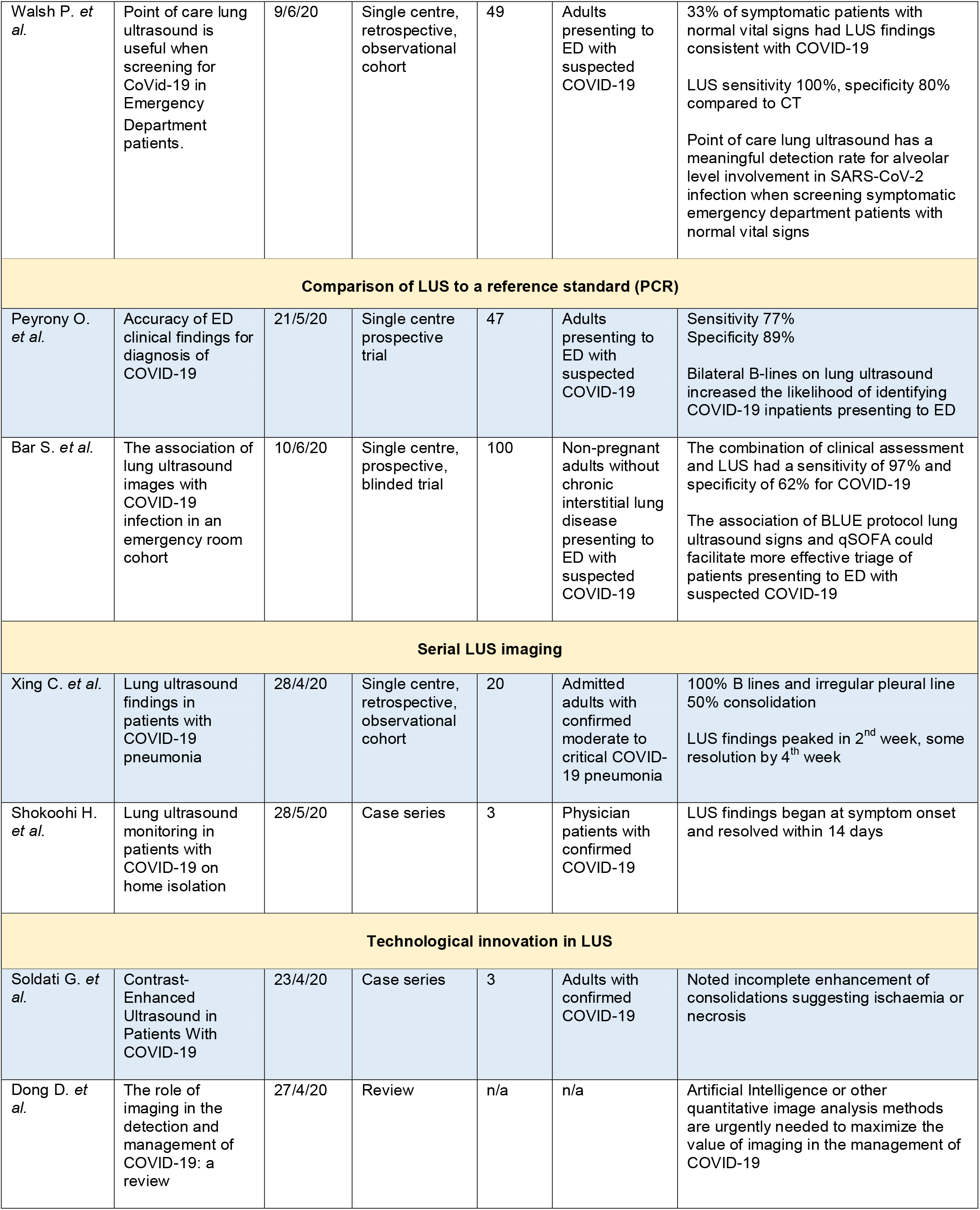
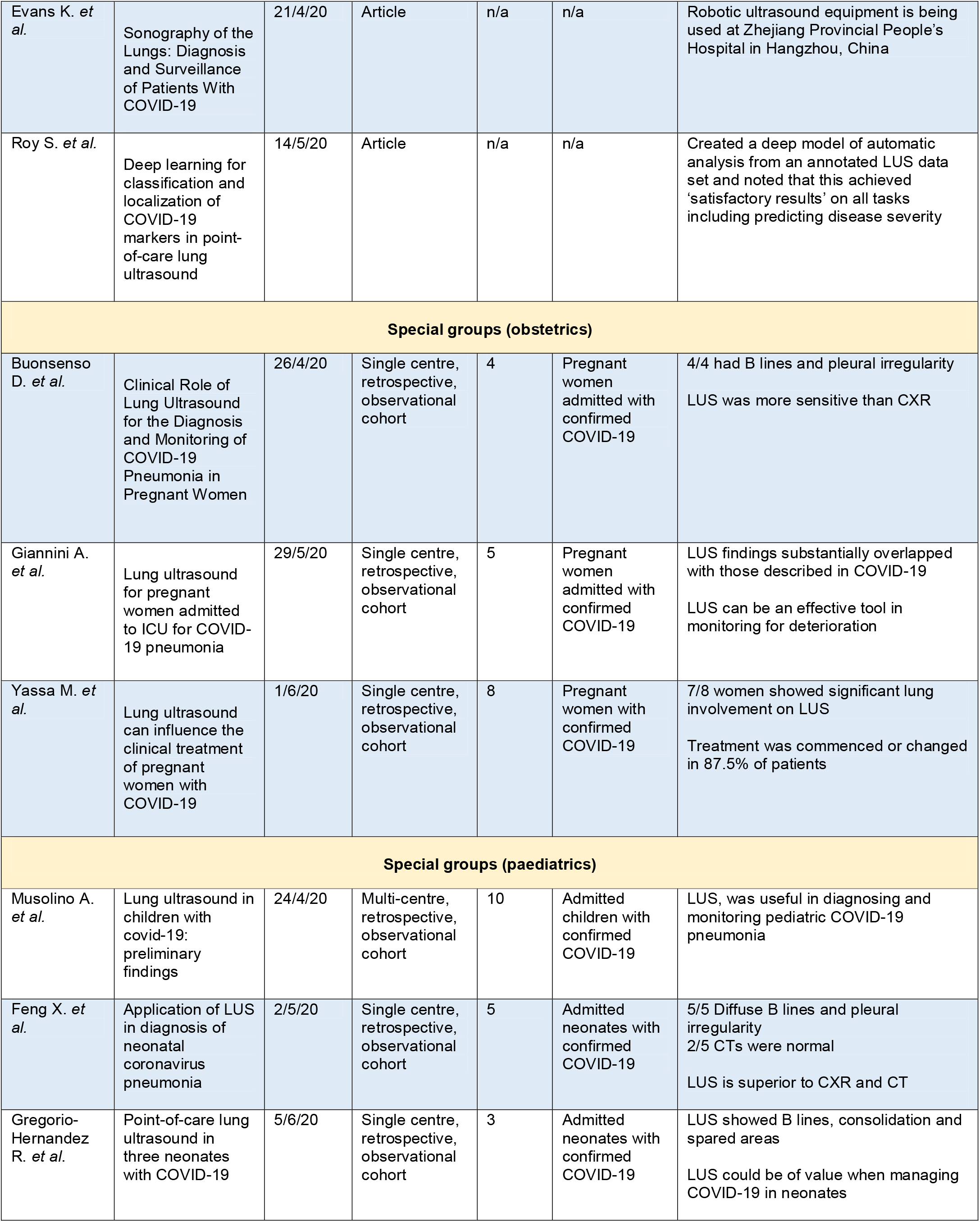
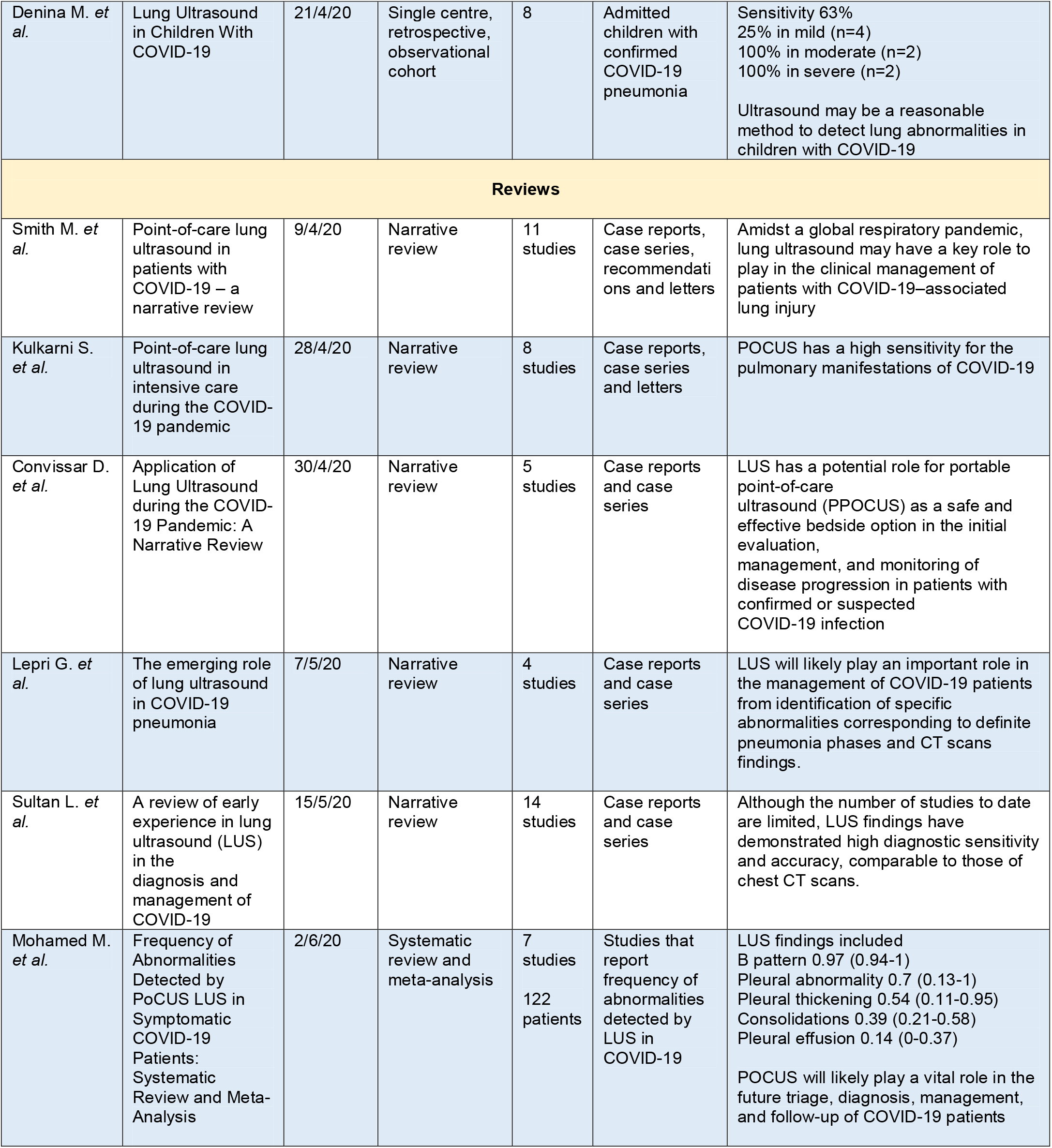

## REFERENCES

1. World Health Organisation. Coronavirus Disease (COVID-19) Dashboard. 2020. https://www.who.int/emergencies/diseases/novel-coronavirus-2019 [Accessed 13 June 2020]

2. Lichtenstein DA, Meziere GA. Relevance of Lung Ultrasound in the Diagnosis of Acute Respiratory Failure: The BLUE Protocol. Chest. 2008; 134:117–25.

3. Volpicelli G, Elbarbary M, Blaivas M, Lichtenstein DA, Mathis G, Kirkpatrick AW, et al. International evidence-based recommendations for point-of-care lung ultrasound. Intensive Care Med. 2012; 38:577–591.

4. Pivetta E, Goffi A, Lupia E, Tizzani M, Porrino G, Ferreri E, et al. Lung-Ultrasound Implemented diagnosis of acute decompensated heart failure in the ED. Chest. 2015; 148:202–210.

5. Orso D, Guglielmo N, Copetti R. Lung Ultrasound in Diagnosing Pneumonia in the Emergency Department: A Systematic Review and Meta-Analysis. Eur J Emerg Med. 2018; 25:312–321.

6. Tsung JW, Kessler DO, Shah VP. Prospective application of clinician-performed lung ultrasonography during the 2009 H1N1 influenza A pandemic: distinguishing viral from bacterial pneumonia. Crit Ultrasound J. 2012; 4:16.

7. Tsai NW, Ngai WC, Mok KL, Tsung JW. Lung ultrasound imaging in avian influenza A (H7N9) respiratory failure. Crit Ultrasound J. 2014; 6:6.

8. Zhang YK, Li J, Yang J, Zhan Y, Chen J. Lung ultrasonography for the diagnosis of 11 patients with acute respiratory distress syndrome due to bird flu H7N9 infection. Virology Journal. 2015; 12:176.

9. Guan W, Ni Z, Hu Y, Liang W, Ou C, He J, et al. Clinical characteristics of coronovirus disease 2019 in China. N Eng J Med. 2020; 382:1708–1720.

10. Wong HYF, Lam HYS, Fong AH, Leung ST, Chin TW, Lo CSY, et al. Frequency and Distribution of Chest Radiographic Findings in COVID-19 Positive Patients. RSNA. EPub ahead of print 27 March 2020. DOI: 10.1148/radiol.2020201160

11. Weinstock MB, Echenique A, Russell JW, Miller JA, Cohen DJ, et al. Chest X-Ray Findings in 636 Ambulatory Patients with COVID-19 Presenting to an Urgent Care Center: A Normal Chest X-Ray Is no Guarantee. J Urgent Care Med. 2020; 14:13–18.

12. Watson J, Whiting PF, Brush JE. Interpreting a COVID-19 test result. BMJ. 2020; 369:m1808.

13. Ai T, Yang Z, Hou H, Zhan C, Chen C, Lv W, et al. Correlation of chest CT and RT-PCR testing in coronavirus disease 2019 (COVID-19) in China: A report of 1014 cases. Radiology. EPub ahead of print 26 Feb 2020. DOI: 10.1148/radiol.2020200642

14. Mongodi S, Orlando A, Arisi E, Tavazzi G, Santangelo E, Caneva L, et al. Ultrasound in patients with acute respiratory failure reduces conventional imaging and healthcare provider exposure to COVID-19. Ultrasound in Med & Biol. EPub ahead of print 6 May 2020. DOI: 10.1016/j.ultrasmedbio.2020.04.033

15. Lu W, Zhang S, Chen B, Chen J, Xian J, Lin Y, et al. A Clinical Study of Noninvasive Assessment of Lung Lesions in Patients with Coronavirus Disease-19 (COVID-19) by Bedside Ultrasound. Ultraschall Med. 2020; 41:300–307.

16. Laursen CB, Sloth E, Lassen AT, Christensen R, Lambrechtsen J, Madsen H, et al. Point-of-care ultrasonography in patients admitted with respiratory symptoms: a single-blind, randomised controlled trial. Lancet. 2014; 2:638–646.

17. Wilkinson J. The final battle – portable ultrasound devices. 2020. https://criticalcarenorthampton.com/2019/04/15/the-final-battle-portable-ultrasound-devices/ [Accessed 1 June 2020]

18. Huang Y, Wang S, Liu Y, Zhang Y, Zheng C, Zheng Y, et al. A preliminary study on the ultrasonic manifestations of peripulmonary lesions of non-critical novel coronavirus pneumonia (COVID-19). SSRN. EPub ahead of print 26 Feb 2020. DOI: 10.2139/ssrn.3544750

19. Soldati G, Smargiassi A, Inchingolo R, Buonsenso D, Perrone T, Briganti DF, et al. Is There a Role for Lung Ultrasound During the COVID-19 Pandemic? J Ultrasound Med. EPub ahead of print 7 April 2020. DOI: 10.1002/jum.15284

20. Moher D, Liberati A, Tetzlaff J, Altman DG. Preferred reporting items for systematic reviews and meta-analyses: The PRISMA Statement. PLOS Medicine. 2020; 6:e1000097.

21. McGowan J, Sampson M, Salzwedel DM, Cogo E, Foerster V, Lefebvre C. PRESS Peer Review of Electronic Search Strategies: 2015 Guideline Statement. J Clin Eidemiol. 2016; 75:40–46.

22. Lomoro P, Verde F, Zerboni F, Simonetti I, Borghi C, Fachinetti C, et al. COVID-19 pneumonia manifestations at the admission on chest ultrasound, radiographs, and CT: single-center study and comprehensive radiologic literature review. Eur J Radiol. 2020; 7:100231.

23. Peng Q, Wang X, Zhang L. Findings of lung ultrasonography of novel corona virus pneumonia during the 2019–2020 epidemic. Intensive Care Med. 2020; 46:849–850.

24. Yasukawa K, Minami T. Point-of-Care Lung Ultrasound Findings in Patients with Novel Coronavirus Disease (COVID-19) Pneumonia. Am J Trop Med Hyg. 2020; 102:1198–1202

25. Volpicelli G, Lamorte A, Villen T. What’s new in lung ultrasound during the COVID-19 pandemic. Intensive Care Med. EPub ahead of print 4 May 2020. DOI: 10.1007/s00134-020-06048-9

26. Mohamed MFH, Al-Shokri S, Yousaf Z, Danjuma M, Parambil J, Mohamed S, et al. Frequency of abnormalities detected by point-of-care lung ultrasound in symptomatic COVID-19 patients: Systematic review and meta-analysis. Am. J. Trop. Med. Hyg. EPub ahead of print 2 June 2020. DOI: 10.4269/ajtmh.20-0371

27. Tung-Chen Y, Marti de Gracia M, DiezTascon A, Agudo-Fernandez S, Alonso-Gonzalez R, Fuertes PR. Correlation between chest computed tomography and lung ultrasonography in patients with coronavirus disease 2019 (COVID-19). MedRxiv. EPub ahead of print 12 May 2020. DOI: 10.1101/2020.05.08.20095117v1

28. Hankins A, Bang H, Walsh P. Point of care lung ultrasound is useful when screening for CoVid-19 in Emergency Department patients. MedRxiv. EPub ahead of print 12 June 2020. DOI: 10.1101/2020.06.09.20123836

29. Yang Y, Huang Y, Gao F, Yuan L, Wang Z. Lung ultrasonography versus chest CT in COVID⍰19 pneumonia: a two⍰centered retrospective comparison study from China. Intensive Care Med. EPub ahead of print 25 May 2020. DOI: 10.1007/s00134-020-06096-1

30. Benchoufi M, Bokobza J, Chauvin AA, Dion E, Baranne M, Levan F, et al. Lung injury in patients with or suspected COVID-19: a comparison between lung ultrasound and chest CT-scanner severity assessments, an observational study. MedRxiv. EPub ahead of print 4 May 2020. DOI: 10.1101/2020.04.24.20069633

31. Lu W, Zhang S, Chen B, Chen J, Xian J, Lin Y. A Clinical Study of Noninvasive Assessment of Lung Lesions in Patients with Coronavirus Disease-19 (COVID-19) by Bedside Ultrasound. Ultraschall in Med. EPub ahead of print 15 April 2020. DOI 10.1055/a-1154-8795

32. Poggiali E, Dacrema A, Bastoni D, Tinelli V, Demichele E, Ramos PM, et al. Can Lung US Help Critical Care Clinicians in the Early Diagnosis of Novel Coronavirus (COVID-19) Pneumonia? Radiology. 2020; 295:3.

33. Lyu G, Zhang Y, Tan G. Transthoracic Ultrasound Evaluation of Pulmonary Changes in COVID-19 Patients During Treatment Using Modified Protocols. Advanced Ultrasound in Diagnosis and Therapy. 2020. 4:79–83.

34. Peyrony O, Marbeuf-Gueye C, Truong V, Giroud M, Riviere C, Khenissi K. Accuracy of Emergency Department clinical findings for diagnostic of coronavirus disease-2019. Ann Emerg Med. EPub ahead of print 21 May 2020. DOI: 10.1016/j.annemergmed.2020.05.022

35. Bar S, Lecourois A, Diouf M, Goldberg E, Bourbon C, Arnaud E, et al. The association of lung ultrasound images with COVID-19 infection in an emergency room cohort. Anaesthesia. EPub ahead of print 10 June 2020. DOI: 10.1111/anae.15175

36. Musolino AM, Supino MC, Buonsenso D, Ferro V, Valentini P, Magistrelli A, et al. Lung Ultrasound in Children with COVID-19: Preliminary Findings. Ultrasound Med Biol. EPub ahead of print. DOI: 10.1016/j.ultrasmedbio.2020.04.026

37. Feng XY, Tao XW, Zeng LK, Wang WQ, Li G. Application of pulmonary ultrasound in the diagnosis of COVID-19 pneumonia in neonates. Zhonghua Er Ke Za Zhi. 2020; 58:347–350.

38. Gregorio-Hernandez R, Escobar-Izquierdo AB, Cobas-Pazos J, Martinez-Gimeno A. Point-of-care lung ultrasound in three neonates with COVID-19. Eur J Paediatr. EPub ahead of print 5 June 2020. DOI: 10.1007/s00431-020-03706-4

39. Denina M, Scolfaro C, Silvestro E, Pruccoli G, Mignone F, Zoppo M, et al. Lung Ultrasound in Children with COVID-19. Paediatrics. EPub ahead of print 1 June 2020. DOI: 10.1542/peds.2020-1157

40. Buonsenso D, Raffaelli F, Tamburrini E, Biasucci DG, Salvi S, Smargiassi A, et al. Clinical role of lung ultrasound for the diagnosis and monitoring of COVID-19 pneumonia in pregnant women. Ultrasound Obstet Gynaecol. EPub ahead of print 26 April 2020. DOI: 10.1002/uog.22055

41. Giannini A, Mantovani A, Vezzoli C, Franchini D, Finazzi P, et al. Lung ultrasound for pregnant women admitted to ICU for COVID-19 pneumonia. Minerva anestesiol. DOI: 10.23736/s0375-9393.20.14726-6

42. Yassa M, Birol P, Mutlu AM, Tekin AB, Sandal K, Tug N. Lung ultrasound can influence the clinical treatment of pregnant women with COVID-19. Journal of ultrasound in medicine EPub ahead of print 1 June 2020. DOI: 10.1002/jum.15367

43. Oxford centre for evidence-based medicine (2009) ‘Levels of evidence’. 2020. https://www.cebm.net/2009/06/oxford-centre-evidence-based-medicine-levels-evidence-march-2009/ [Accessed 1 June 2020]

44. Rippey J. Lung ultrasound: comprehensive examination. 2018. https://litfl.com/lung-ultrasound-comprehensive-examination/ [Accessed 1 June 2020]

45. Cox EGM, Wiersema R, Wong A, Van Der Horst ICC. Six versus eight and twenty-eight scan sites for B-line assessment: differences in examination time and findings. Intensive Care Med. 2020; 46:1063–1064.

46. Volpicelli G, Gargani L. Sonographic signs and patterns of COVID-19 pneumonia. The Ultrasound Journal. 2020; 12:22.

47. Soldati G, Smargiassi A, Inchingolo R, Buonsenso D, Perrone T, Briganti DF, et al. On lung ultrasound patterns specificity in the management of COVID-19 patients. J Ultrasound Med. 2020; 9999:1–2.

48. Pivetta E, Goffi A, Nazerian P, Castagno D, Tozzetti C, Tizzani P, et al. Lung ultrasound integrated with clinical assessment for the diagnosis of acute decompensated heart failure in the emergency department: a randomized controlled trial. European Journal of Heart Failure. 2019; 21:754–766.

49. Leeflang MMG, Rutjes AWS, Reitsma JB, Hooft L, Bossuyt PMM, et al. Variation of a test’s sensitivity and specificity with disease prevalence. CMAJ. 2013; 185:537–544.

50. Xing C, Li Q, Du H, Kang W, Lian J, Yuan L. Lung ultrasound findings in patients with COVID-19 pneumonia. Critical Care. 2020; 24:174.

51. Shokoohi H, Duggan NM, Sanchez GG, Torres-Arrese M, Tung-Chen Y. Lung ultrasound monitoring in patients with COVID-19 on home isolation. American Journal of Emergency Medicine. EPub ahead of print 28 May 2020. DOI: 10.1016/j.ajem.2020.05.079

52. Rouby JJ, Arbelot C, Gao Y, Zhang M, Lv J, An Y, et al. Training for lung ultrasound score measurement in critically ill patients. Am J Respir Crit Care Med. 2018; 198:398–401.

53. Dong D, Tang Z, Wang S, Hui H, Gong L, Lu Y, et al. The role of imaging in the detection and management of COVID-19: a review. IEEE. EPub ahead of print 27 April 2020. DOI: 10.1109/RBME.2020.2990959

54. Roy S, Menapace W, Oei S, Luitjen B, Fini E, Saltori C. Deep learning for classification and localization of COVID-19 markers in point-of-care lung ultrasound. IEEE. EPub ahead of print 14 May 2020. DOI: 10.1109/TMI.2020.2994459

55. Evans KD, Yang Q, Liu Y. Sonography of the Lungs: Diagnosis and Surveillance of Patients With COVID-19. Journal of Diagnostic Medical Sonography. EPub ahead of print 21 April 2020. DOI: 10.1177/8756479320917107

56. Soldati G, Giannasi G, Smargiassi A, Inchingolo R, Demi L. Contrast-Enhanced Ultrasound in Patients With COVID -19. J Ultrasound Med. EPub ahead of print 12 May 2020. DOI: 10.1002/jum.15338

